# Phenotyping of Indian Polycystic Ovarian Syndrome on the basis of Biochemical Parameters: A retrospective analysis via Hierarchical Cluster Classification

**DOI:** 10.1101/2020.11.16.20232942

**Authors:** Janani Dakshina Moorthy, Rajapriya Ayyappan, B Usha

## Abstract

**Objective:** We aimed to phenotype the Indian PCOS population based on their etiology for an effective treatment regimen.

**Design:** Retrospective analysis of biochemical data.

**Setting:** PCOS clinics in Tamil Nadu, India

**Population or Sample:** Girls and women in age group 18 to 30 diagnosed as PCOS by RC.

**Method:** The statistical analysis was done using two-way cluster analysis function of SPSS v.22 to identify the phenotypes and the resolving biochemical parameter. Also, the population was segregated into three cohorts based on their age for further analysis.

**Main outcome measure:** Endocrine parameters like LH, FSH, estradiol, testosterone and thyroid profile. Biochemical parameters like complete lipid profile, blood glucose and insulin fasting. Body Mass Index (BMI).

**Results:** The statistical analysis reported two phenotypes among the Indian PCOS population, segregated based on their LH: FSH ratio. The phenotype with LH: FSH >2, had a hormonal imbalance and may have its etiology from Hypothalamus – Pituitary - Ovarian axis. The phenotype with LH: FSH < 2 had significant markers indicating the incidence of metabolic syndrome and may follow an insulin – dependent pathway for PCOS manifestation.

**Conclusion:** The PCOS population needs a comprehensive screening before deciding on a treatment regimen. All the PCOS patients need to be recommended to follow an active lifestyle since 80% of them are predisposed to a metabolic syndrome in their later ages.

## Introduction

Polycystic Ovarian Syndrome (PCOS) is the most common gynaecological endocrinology disorder affecting adolescent girls to older women, being evident among reproductive women [1]. Indian PCOS clinics employ Rotterdam criteria 2003 (RC), for diagnosis, as recommended by the Indian Fertility Society (IFS) with some evidence-based modification for diagnosing Indian PCOS [2]. Subsequently, the IFS guidelines also insisted the documentation of specific Indian phenotypes identified upon diagnosis and carry out research for assessing the adaptation of RC specific to the Indian ethnicity [3].

Until now researchers in India have documented PCOS phenotypes based on RC. They have identified four phenotypes using the diagnosis criteria Hyperandrogenism (H), Oligo anovulation (O) and Polycystic Ovaries (P). The phenotypes identified are (i) H + P (ii) P + O (iii) H + O (iv) H + P + O. These phenotypes are based on the symptoms rather than the etiology [4-6].

India is also identified to be a “severe metabolic” PCOS phenotype and subsequently a few studies from India have correlated the RC based PCOS phenotypes and prevalence of metabolic syndrome among Indians [5-7]. Though a couple of these studies provide evidence that hyperandrogenic PCOS phenotype is more susceptible to a metabolic syndrome [8,9]; they fail to include pathological parameters and thus does not give clarity on treatment.

Current treatment protocols in India are designed to alleviate the cosmetic effects of hyperandrogenism and insulin resistance, regularize the menstruation, and manage infertility. According to IFS guidelines, the Combined Oral Contraceptives (COCs) remain the first-line treatment even though there is evidence for the incidence of metabolic syndrome. Insulin sensitizers are prescribed only if COCs fail to restore cycle and if the patient is trying to conceive even though the patient shows no obvious Insulin Resistant symptoms[3]. Thus, the treatment for PCOS is usually a trial and error protocol. Efficient management of PCOS potentially by attending to its pathology may reduce the risk of such complications.

However, tailoring the therapy according to the needs of an individual patient pertaining to the varied manifestations of PCOS can be a complex clinical implementation. Moreover, the diagnosis of PCOS symptoms and its associated morbidities itself proves challenging and needs a standard protocol.

To overcome this complication in the diagnosis and for a personalized treatment, we propose a strategy to group Indian PCOS population based on their biochemical profiles. This grouping, unlike RC, will categorize the PCOS patients based on the pathology rather than the symptoms.

## Methods

### Data collection

This is a retrospective analysis of the data available from a couple of tertiary care units during a time frame spanning one year (April 2017 and April 2018). The biochemical reports of 250 PCOS patients fitting the inclusion criteria were collected. The reports were anonymized the minute they were obtained and the information was gathered and tabulated into a computerized version for automation of analysis.

The inclusion criteria for choosing a biochemical report were: (i) The report should be of a patient diagnosed as PCOS under the RC [10] (ii) The report must carry information about endocrine parameters (iii) the report should encompass the markers for diagnosing metabolic syndrome according to the National Cholesterol Education Program (NCEP) Adult Treatment Panel (ATP) III guidelines [11].

### Comparing the Biochemical Evaluation

To minimize errors a few critical points were taken into consideration as the reports were collected from two centres. The routine blood sampling should have been taken during the early follicular phase, between days 2 and 3 from the last menstrual period (LMP) to obtain the basal values. For women with PCOS who do not undergo normal menstruation, the blood sampling should be performed during the follicular phase of an induced cycle.

Though the sample collection, handling, and processing were done following the internal laboratory standards and GLP, the assay methods and reporting format for each parameter were ensured to be in identical format. The HOMA – IR 1 scores for every individual was calculated using serum insulin and glucose levels reported in their respective biochemistry report. The reference range is as recommended by the Food and Drug Administration (FDA).

### Statistical Analysis

The Statistical Packages for Social Sciences, SPSS version 22 (SPSS, Chicago, IL, USA), was used for data analysis. Continuous variables are presented as mean and standard deviation where as categorical variables are expressed in percentage. Baseline characteristics are presented as a mean ± standard deviation and the differences between groups were assessed by using chi square-test for categorical variables. The statistically significance was calculated at 95% confidence interval.

In order to categorize data based on the phenotypes of PCOS from the available biochemical parameters, we carried out a two-step cluster analysis. The grouping was performed by running pre-clustering first and then by hierarchical methods. This hierarchy was represented as a dendrogram for visual perception.

For a comprehensive analysis, we had partitioned the biochemical data into three categories based on their age (Table 1.a and Table 1.b, Supplementary Table 2 &3). The first category had the pubertal and postpubertal age group which includes ages from 13 years to 18 years. The second category included ages from 19 to 25, which is an active reproductive age and the third group included the ages of 26 to 30.

**Table 1.a.** Hormonal attributes of the cohorts HI and MetS

|  | Hormonal Imbalance Phenotype Group |  |  |  |  | Metabolic Syndrome Phenotype Group |  |  |  |  |
| --- | --- | --- | --- | --- | --- | --- | --- | --- | --- | --- |
| Age Group | Testosterone | LH | FSH | Estradiol | TSH | Testosterone | LH | FSH | Estradiol | TSH |
| 13 - 18 | 0.54±0.14 | 12.24±8.1 | 4.19±1.7 | 114.28±64.1 | 4.62±0.6 | 0.98±0.34 | 3.67±2.27 | 4.49±1.03 | 41.09±13.6 | 2.73±1.67 |
| 19 - 25 | 0.63±0.04 | 11.62±7.2 | 3.82±2.0 | 159.9±145.7 | 6.21±14.7 | 1.14±0.94 | 4.2±2.09 | 4.31±1.8 | 65.9±39.6 | 2.88±1.69 |
| 25 - 30 | 0.54±0.1 | 10.13±6.4 | 3.45±2.1 | 231.28±32.1 | 3.42±3.2 | 0.42±0.06 | 4.2±2.09 | 4.31±1.8 | 70.41±49.08 | 2.88±1.69 |

## Results

Out of the 250 reports considered for the study, three were rejected due to incomplete data and thus only 247 reports were analyzed.

### Cluster Analysis Illustrated the Prevalence of Two Indian PCOS Phenotype Based on Pathology

The cluster analysis on biochemical data, sorted the study cohort into two groups of patients that present with similar clinical symptoms but have a different biochemical diagnosis (Fig. 1). The first group was constitutted with hormonal features like LH, FSH, and Thyroid. This group was found to represent the characteristics of a typical PCOS hormonal disturbance and is labeled as Hormonal Imbalance Group (HI) for the purpose of this study. The other group consisted of benchmarked hormonal parameters indicating the mpossible occurrence of metabolic syndrome and is labeled as Metabolic Syndrome Phenotype group (MetS).

**Fig. 1.a.**
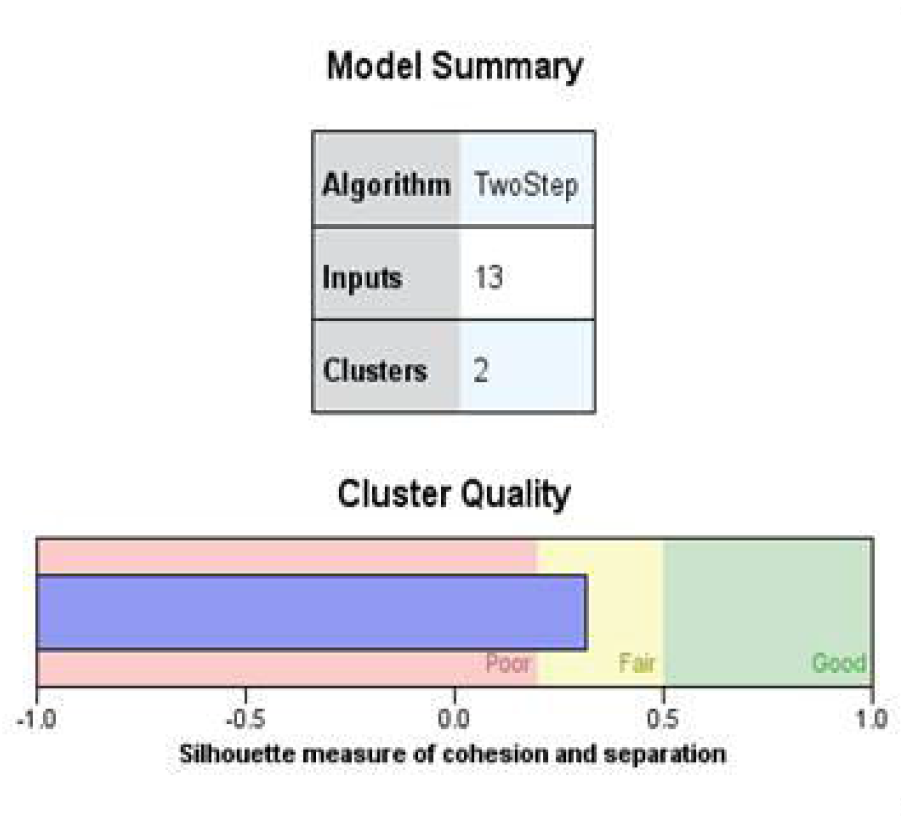
Model summary of the two-step cluster analysis with a graph of the quality of clusters.

**Fig. 1.b.**
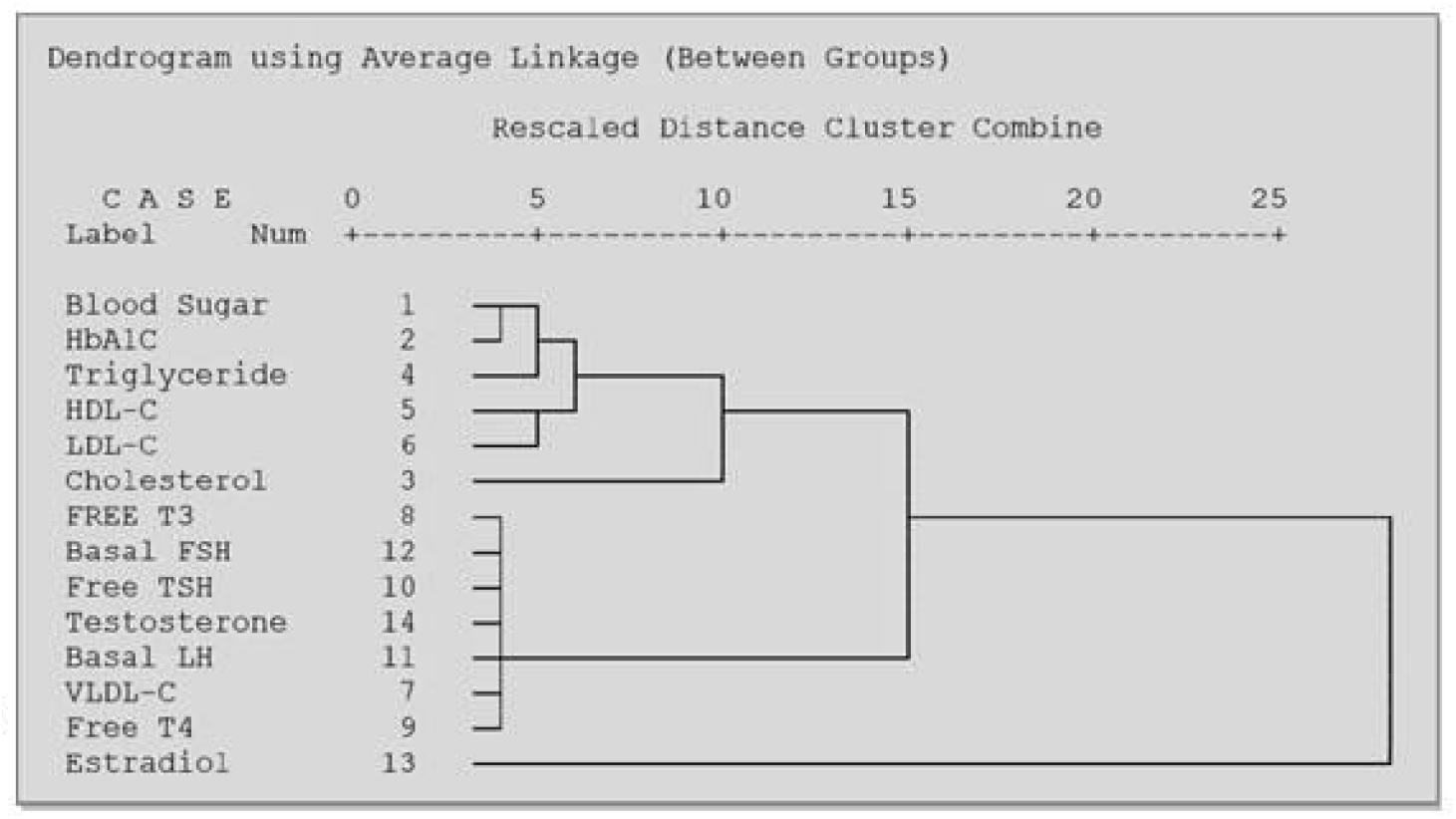
Hierarchical representation of the diagnostic parameters for the two PCOS Phenotypes.

Prematurely before any PCOS diagnosis criteria were established for clinical practise, LH: FSH ratio was considered as a golden standard clinical surrogate marker where patients with a ratio > 2 were diagnoised as PCOS [12]. Also from our statistical analysis, it was observed that LH demonstrated an influence on both the groups, as represented in the dendrogram (SF1). Thus, we applied the LH: FSH ratio with a threshold of 2, as the determining parameter to assign a biochemical profile to either of the clusters – HI or MetS. The clusters furnished the distinct and comprehensive characteristics of the phenotypes forming the groups as described in Supplementary Table 1(ST1).

We found that majority of the Indian ethnic PCOS women fall under the MetS category (78.4%) and only one fourth of the population have endocrinal disruptions (21.6%). Thus when the groups were segregated to analyze for each trait, discrimination in the number of variables among the two groups was found. Thus an additional statistical significance was also calculated to ensure that results are not biased by group size and reported in ST1.

### Hormonal Attributes of the HI and MetS Phenotypes

The traits considered for hormonal analysis were LH, FSH, complete thyroid profile, Estradiol and Testosterone. A prominent finding was that the LH concentration was considerably higher than basal values for HI cohort and was almost thrice their MetS counterpart (HI = 11.39 ± 7.37; MetS = 4.2 ± 2.4; p* < 0.0001). The serum TSH levels of the HI cohort was in proximity to the higher end of the normal range and thus demonstrated an inclination for development of thyroid disorder; while the MetS group expressed no such indications (HI = 4.99 ± 11.2; MetS = 3.2 ± 2.6; p* < 0.01). The estradiol (E2) concentration was almost three-fold higher than basal value in the HI group (HI = 167.4 ± 200.4; MetS = 64.6 ± 43.06; p* < 0.001). The serum testosterone levels (T), an important PCOS diagnostic marker, was found to be at borderline for HI cohort (HI = 0.7 ± 0.33; MetS = 1.08 ± 1.8; p* <0.005) and is found to be elevated among the MetS cohort. This indicates that HI cohort may not be hyperandrogenic but may have their pathophysiology from other hormones especially LH.

The hormonal variations as the age progresses among both the cohorts are detailed in Table 1.a. An evident characteristic of this categorizing was that the LH: FSH ratio tends to become normal as age progressed i.e younger cohorts had a defined LH: FSH ratio > 2 and the oldest group had LH: FSH ratio almost equal to 1. This finding adds evidence to the claim that PCOS initially represents as a hormonal disorder and manifests into a metabolic syndrome with aging, thus highlighting the importance of long term follow – up of PCOS women.

### Metabolic Aspects of HP and MetS Phenotypes

The BMI of the MetS cohort was found to be alarmingly high (HI = 28.7 ± 0.8; MetS = 33.4± 1.2; p* <0.0001) in comparison with the HI group; however, on the whole the BMI was found to be higher than the ideal range among Indian PCOS women. The total cholesterol (HI = 147.8 ± 36.1; MetS = 155.43 ± 27.9; p* < 0.001) and triglycerides (HI = 94.11 ± 34.3; MetS = 99.3 ± 3; p* < 0.005) were comparatively high in the MetS cohort and the High Density Lipoprotein – Cholesterol (HDL – C) was found to be below the required concentration (HI = 46 ± 10; MetS = 44.13 ± 9.9; p* <0.005) in the same group, indicating strong tendency towards development of metabolic syndrome. The HDL – C serum concentration was also found to be lower than the essential amount for its protective effect on the HI cohort. The magnified serum Testosterone (T) levels along with aberrant HDL – C and E2 levels are found to be characteristic to the MetS, which may eventually lead to development of a metabolic syndrome in later stages of life.

In an extended analysis, we found that among the study cohort 146 PCOS candidates were having very low HDL concentrations and the conditions seem hazardous especially for the 10.6% (N = 15/146; M: 176.5 ± 7.5; CI: 139.6 - 213.3) of this cohort, as they also had high levels of triglycerides and thus prone to cardiovascular disorders at early stages of life.

The incidence of metabolic syndrome symptoms across various age groups is depicted in Table 1.b. A crucial finding of the age – wise analysis was that the youngest age group, i.e 13 to 18 year, had elevated triglyceride concentration and HDL concentration was below the protective levels. Also it must be noticed that the estradiol concentration was comparably low for this cohort and had the highest BMI range. These findings highlight the predisposition of these adolescent women towards developing metabolic syndromes like obesity and cardiovascular disorder prematurely.

**Table 1.b.**
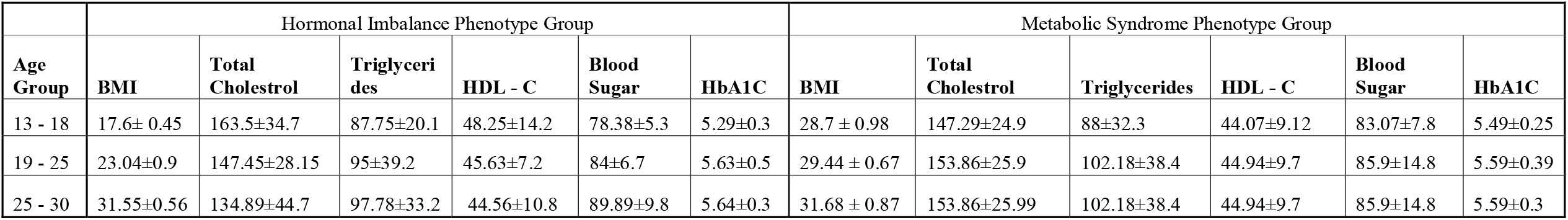
Metabolic attributes of the cohorts HI and MetS

## Discussion

### Main Findings

Epidemiological studies have revealed that the PCOS prevalence and manifestation vary among ethnic regions [13]. India is a country with the most diverse ethnicity having distinguished health patterns [14] and the high occurrence of PCOS (1 out of every 6 women) [15]. Thus, PCOS is very diversely presented among Indian women and these differences in presentation of the syndrome may possibly be due to deviant inclusion criteria, unrepresentative clinical manifestation and metabolically varying impacts of the syndrome.

This massive disorder challenges the practitioners in terms of diagnosis and therapy due to its complex nature of the PCOS. However, it is supposed by the IFS that if a preliminary risk assessment is stratified consensus to an Indian clinical practice, further referrals to tertiary care units will be more defined. This stratification may not be an alternative to the diagnosis criteria but will work in coordination, for systemic diagnosis and treatment regimen [3].

Consequently, study subjects from India are clustered into two groups, HI and MetS phenotypes, based on their diagnostic biochemical traits and the LH: FSH ratio was found to serve as the resolving parameter. HI cohort tends to have an endocrine dysfunction and MetS appear to have metabolic dysfunction as their pathophysiology.

### Strength

This is the first study which aimed to group the Indian PCOS population into distinct phenotypes based on biochemical aspects. The strength of the study lies in the fact that it includes patients from most of the Indian ethnicity. Since the grouping was based on the basal serum biochemical and endocrinological parameters rather than the RC, which screens for the symptoms rather than the etiology; the treatment following the grouping pattern will address the root cause of the disorder instead of remedy for symptoms.

### Limitations

The drawback in the study was that it was not a prospective analysis to strategically analyze for specific parameters and also the patients were not sorted into ethnic groups. Also the pattern for the biochemical parameters of age matched controls need to be observed.

### Interpretation

Current literatures are suggestive of two pathways for the pathology of PCOS. One is based on Hyperinsulinemia [16] and the other is due to a disruption in the Hypothalamus-Pituitary – Ovary (HPO) axis [17]. The hyperinsulinemia condition induces steroidogenesis in thecal cells mediated by the activation of PI3K pathway causing a decreased phosphorylation of MEK/ERK signaling, which results in increased CYP17 gene expression and androgen biosynthesis (Fig. 2) [18 – 22]. The HPO axis is disrupted when the hypothalamus and pituitary gland becomes insensitive to the negative feedback mechanism of LH caused by the chronic exposure of hypothalamus to estrogen (Fig. 3) [23 – 26].

**Fig. 2.**
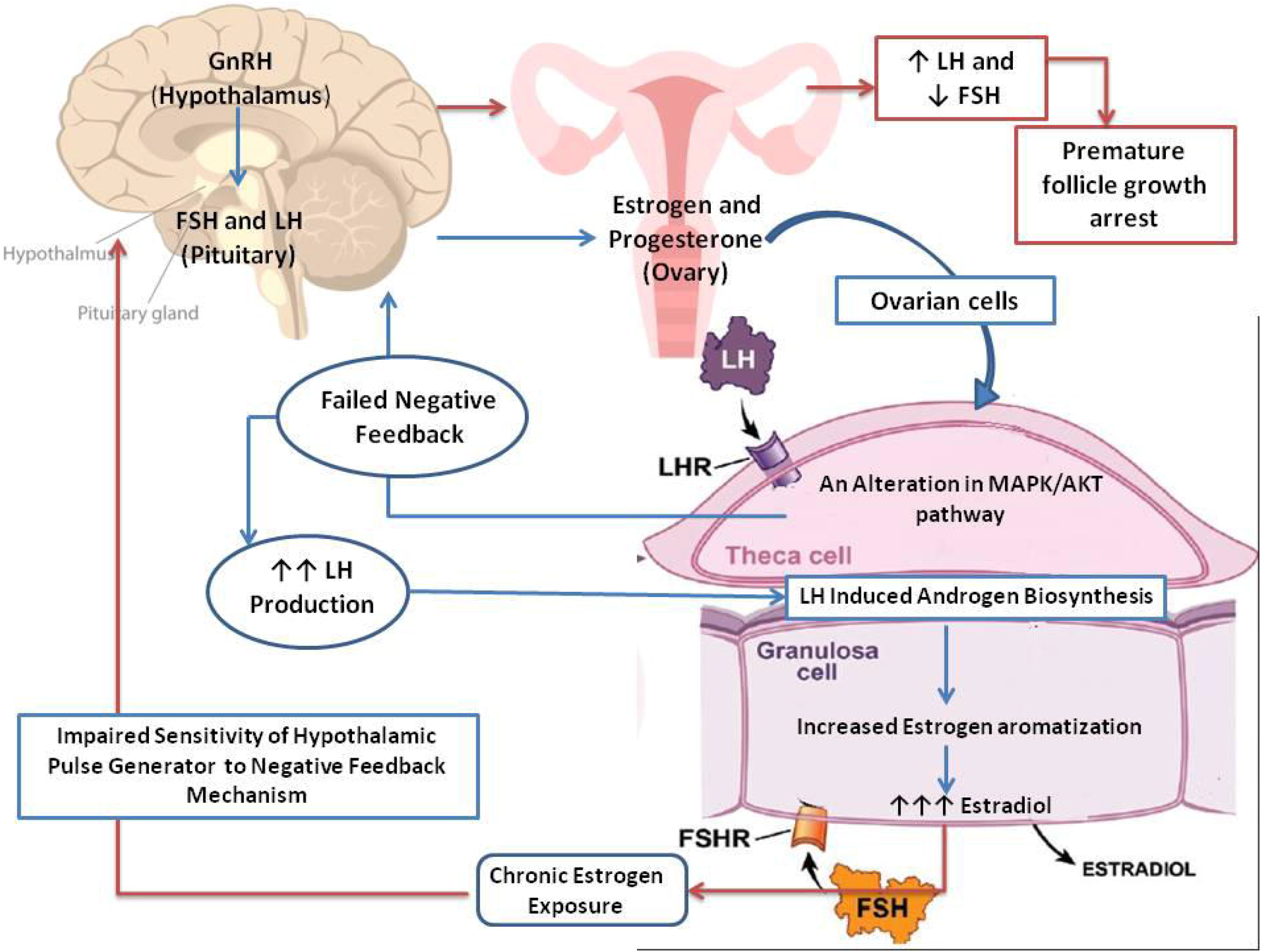
Pathology of PCOS for Hormonal Imbalance Phenotype (HI) Cohort.

**Fig. 3.**
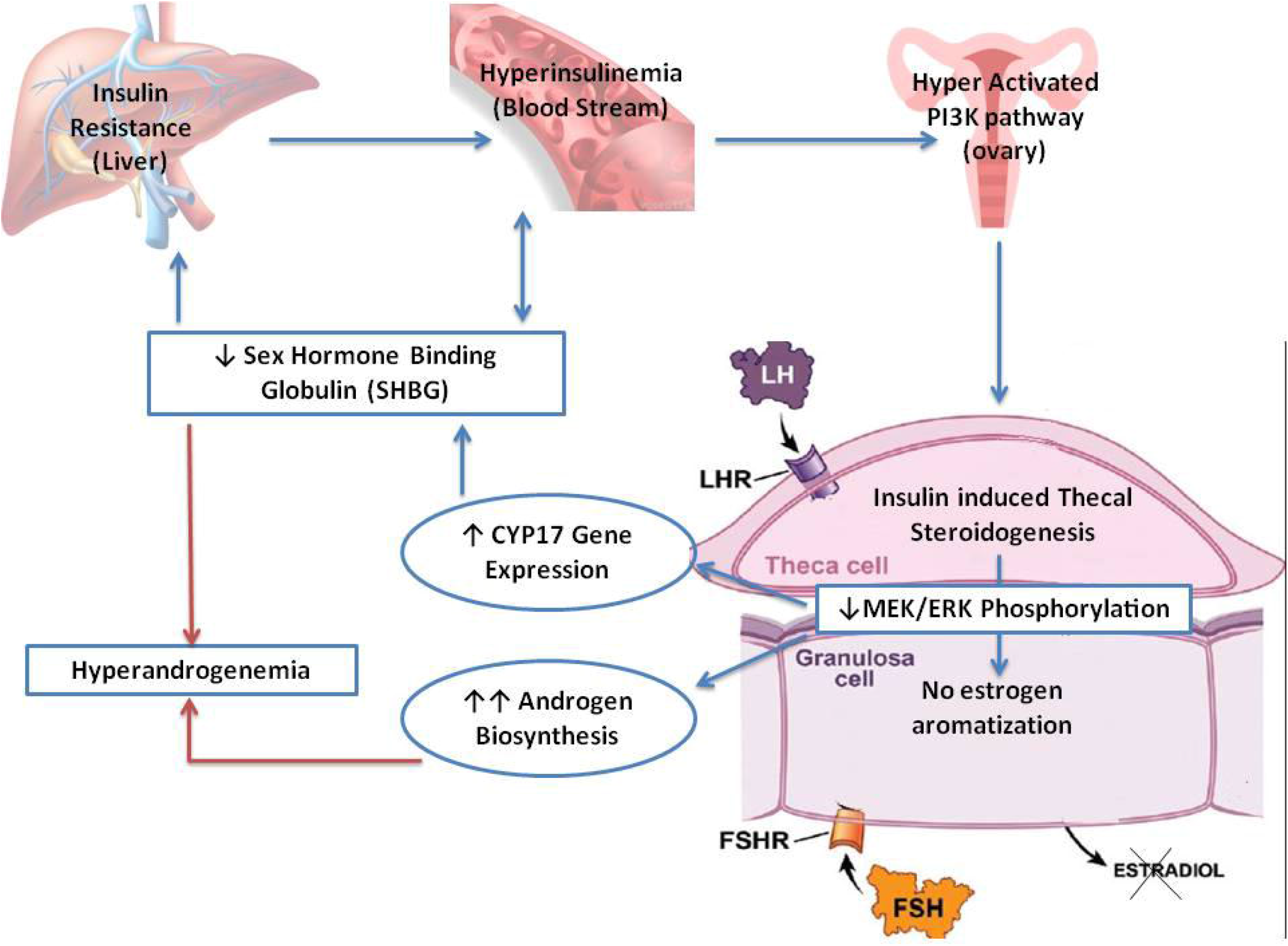
Pathophysiology of hyperinsulinemia leading to PCOS in Metabolic Syndrome Phenotype (MetS) group.

Based on our hypothesis, the cohort under Hormonal Imbalance phenotype (HI) show endocrine disturbances and has notably elevated concentrations of luteinizing hormone (LH) while their serum glucose levels do not appear to be disturbed. Thus, we contemplate the HI group to have a pathophysiology which is insulin independent and have disturbances in the HPO axis. Thus, it is not rationale to prescribe insulin sensitizers or lifestyle modifications as first-line treatment for this phenotype. An appropriate remedy would be Combined Oral Contraceptives (COCs) which control the androgen steroidogenesis and enhances the bioelimination of androgens by enhancing the production of Sex Hormone Binding Globulin (SHBG) by the liver [27]. Thus COCs may be administered for conquering menstrual irregularities for the HP after complete clinical and biochemical investigation. A careful and comprehensive screening is important to decide upon the type and dosage of COCs to be administered. COCs cannot be taken for a long duration because of its adverse side effects and menstrual irregularities seem to reappear after withdrawal of medication [28,29]. Thus a cure yet needs to be devised for this phenotype.

The Metabolic Syndrome Group (MetS) clearly are indicative of a predisposition to cardiovascular disorders (CVD) and type II Diabetes Mellitus [30,31]. Also in this cohort, a notable factor is that the hormones LH, T and E2 in the serum remain within the basal value and thus satisfy the anovulation and polycystic ovary (PCO) criteria of the RC. Since the PCO condition and anovulation go hand in hand, the PCOS status of these candidates needs to be reconsidered by contemplating biochemical markers with clinical symptoms [32]. Women under this cohort may be treated with insulin sensitizers and may be recommended lifestyle modifications for early sensitizing of long term consequences. Insulin sensitizers are known to reduce the circulating insulin levels, enhance the normal proliferation of theca cells and thus regulate the androgen biosynthesis; thus alleviating the cosmetic symptoms, which is an attribute of this phenotype and improves the endometrial growth thus initiating the regular menstrual cycle. And also this group needs to be further evaluated and channeled to the appropriate tertiary care unit for long-term follow up in order to prevent or manage the associated morbid conditions.

But the RCOG guidelines, recommend the administration of insulin sensitizers only to those individuals with low glucose tolerance and the IFS recommends following international guidelines while treating for PCOS. Considering the “severe metabolic” phenotype of Indian PCOS population and the possibility of hyperinsulinemia to manifest itself in the form of obesity or fat deposition, insulin sensitizers may confer relief from the metabolic syndrome symptoms [33].

Thus as hypothesized, placing a PCOS individual seeking medical help under either of the phenotypes as identified by cluster analysis may help the clinician by giving an insight into the pathophysiology of the individual and also to decide upon a treatment regimen. The LH: FSH ratio from a patient’s biochemical profile will help the clinician assign the individual to either the HI or MetS group and thus be treated appropriately.

Therefore we recommend that when a woman is being treated for effects of PCOS like the irregularity of cycles, acne, facial hair or infertility, the manifestations of PCOS in the long run and its metabolic syndrome prevalence should also be given corresponding importance and guarded. This study also suggests that the diagnosis regimen, treatment module, and long-term follow - up for each phenotype should be managed distinctly.

## CONCLUSION

Though the LH: FSH ratio has remained the gold standard for diagnosing PCOS, this rule did not directly imply as a diagnosing criterion for the Indian population, who probably have a different phenotype from other ethnic populations. But this ratio seems to be the resolving factor in categorizing the Indian PCOS population for deciding a treatment regimen.

The results of our study indicate the need for a methodical analysis of biochemical parameters and clinical investigations, to make the treatment module absolute to overcome complexities such as the lack of standard diagnostic criteria. We recommend a comprehensive screening of ovulatory patterns, hirsutism, all the endocrinological factors, biochemical parameters, and metabolic syndrome prevalence before affirming the PCOS status and starting with a first line medication based on the etiology rather than simply alleviating the clinical symptoms. Also, it is essential to educate the women of all ages with PCOS of their long-term risk of type 2 Diabetes and likely cardiovascular disease and the importance of constant monitoring.

## Data Availability

The complete data has been recoded in the format of a Microsoft Excel spreadsheet.

## References

1. Baldani DP, Skrgatic L, Ougouag R. Polycystic ovary syndrome: important underrecognised cardiometabolic risk factor in reproductive-age women. International journal of endocrinology. 2015;2015.

2. Legro RS. Evaluation and Treatment of Polycystic Ovary Syndrome. Available from: https://www.ncbi.nlm.nih.gov/books/NBK278959/ accessed on October 23, 2018.

3. Malik S, Jain K, Talwar P, Prasad S, Dhorepatil B, Devi G, Khurana A, Bhatia V, Chandiok N, Kriplani A, Shah D. Management of polycystic ovary syndrome in India. Fertility Science and Research. 2014 Jan 1;1(1):23.

4. Ramanand SJ, Ghongane BB, Ramanand JB, Patwardhan MH, Ghanghas RR, Jain SS. Clinical characteristics of polycystic ovary syndrome in Indian women. Indian journal of endocrinology and metabolism. 2013 Jan;17(1):138.

5. Mandrelle K, Kamath MS, Bondu DJ, Chandy A, Aleyamma TK, George K. Prevalence of metabolic syndrome in women with polycystic ovary syndrome attending an infertility clinic in a tertiary care hospital in south India. Journal of human reproductive sciences. 2012 Jan;5(1):26.

6. Sobti S, Dewan R, Ranga S. Metabolic syndrome and insulin resistance in PCOS phenotypes. International Journal of Reproduction, Contraception, Obstetrics and Gynecology. 2017 Oct 28;6(11):5067–73.

7. Casarini L, Brigante G. The polycystic ovary syndrome evolutionary paradox: a genome-wide association studies–based, in silico, evolutionary explanation. The Journal of Clinical Endocrinology & Metabolism. 2014 Nov 1;99(11):E2412–20.

8. Tripathy P, Sahu A, Sahu M, Nagy A. Metabolic risk assessment of Indian women with polycystic ovarian syndrome in relation to four Rotterdam criteria based phenotypes. European Journal of Obstetrics & Gynecology and Reproductive Biology. 2018 May 1;224:60–5.

9. Kar S. Anthropometric, clinical, and metabolic comparisons of the four Rotterdam PCOS phenotypes: A prospective study of PCOS women. Journal of human reproductive sciences. 2013 Jul;6(3):194.

10. Fr DD, Tarlatzis R. Revised 2003 consensus on diagnostic criteria and long-term health risks related to polycystic ovary syndrome. Fertility and sterility. 2004 Jan;81(1).

11. Stone NJ, Bilek S, Rosenbaum S. Recent national cholesterol education program adult treatment panel III update: adjustments and options. The American journal of cardiology. 2005 Aug 22;96(4):53–9.

12. Homburg R. What is polycystic ovarian syndrome? A proposal for a consensus on the definition and diagnosis of polycystic ovarian syndrome. Human Reproduction. 2002 Oct 1;17(10):2495–9.

13. Goldrat O, Van Den Steen G, Gonzalez-Merino E, Dechène J, Gervy C, Delbaere A, Devreker F, De Maertelaer V, Demeestere I. Letrozole-associated controlled ovarian hyperstimulation in breast cancer patients versus conventional controlled ovarian hyperstimulation in infertile patients: assessment of oocyte quality related biomarkers. Reproductive Biology and Endocrinology. 2019 Dec;17(1):3.

14. Mungreiphy NK, Dhall M, Tyagi R, Saluja K, Kumar A, Tungdim MG, Sinha R, Rongmei KS, Tandon K, Bhardwaj S, Kapoor AK. Ethnicity, obesity and health pattern among Indian population. Journal of natural science, biology, and medicine. 2012 Jan;3(1):52.

15. Bharathi RV, Swetha S, Neerajaa J, Madhavica JV, Janani DM, Rekha SN, Ramya S, Usha B. An epidemiological survey: Effect of predisposing factors for PCOS in Indian urban and rural population. Middle East Fertility Society Journal. 2017 Dec 1;22(4):313–6.

16. Reddy BM, Kommoju UJ, Dasgupta S, Rayabarapu P. Association of type 2 diabetes mellitus genes in polycystic ovary syndrome aetiology among women from southern India. The Indian journal of medical research. 2016 Sep;144(3):400.

17. Allahbadia GN, Merchant R. Polycystic ovary syndrome and impact on health. Middle East Fertility Society Journal. 2011 Mar 1;16(1):19–37.

18. Li T, Mo H, Chen W, Li L, Xiao Y, Zhang J et al. Role of the PI3K-Akt signaling pathway in the pathogenesis of polycystic ovary syndrome. Reproductive sciences. 2017 May;24(5):646–55.

19. Liu L, Kang J, Ding X, Chen D, Zhou Y, Ma H. Dehydroepiandrosterone-regulated testosterone biosynthesis via activation of the ERK1/2 signaling pathway in primary rat leydig cells. Cellular Physiology and Biochemistry. 2015;36(5):1778–92.

20. Udhane SS, Flück CE. Regulation of human (adrenal) androgen biosynthesis—New insights from novel throughput technology studies. Biochemical pharmacology. 2016 Feb 15;102:20–33.

21. Banerjee U, Dasgupta A, Khan A, Ghosh MK, Roy P, Rout JK, et al. A cross-sectional study to assess any possible linkage of C/T polymorphism in CYP17A1 gene with insulin resistance in non-obese women with polycystic ovarian syndrome. The Indian journal of medical research. 2016 Jun;143(6):739.

22. Winters SJ, Gogineni J, Karegar M, Scoggins C, Wunderlich CA, Baumgartner R, Ghooray DT. Sex hormone-binding globulin gene expression and insulin resistance. The Journal of Clinical Endocrinology & Metabolism. 2014 Dec 1;99(12):E2780–8.

23. Thompson IR, Kaiser UB. GnRH pulse frequency-dependent differential regulation of LH and FSH gene expression. Molecular and cellular endocrinology. 2014 Mar 25;385(1-2):28–35.

24. Makker A, Goel MM, Das V, Agarwal A. PI3K-Akt-mTOR and MAPK signaling pathways in polycystic ovarian syndrome, uterine leiomyomas and endometriosis: an update. Gynecological Endocrinology. 2012 Mar 1;28(3):175–81.

25. Bliss SP, Navratil AM, Xie J, Roberson MS. GnRH signaling, the gonadotrope and endocrine control of fertility. Frontiers in neuroendocrinology. 2010 Jul 1;31(3):322–40.

26. Lagrange AH, Rønnekleiv OK, Kelly MJ. Estradiol-17 beta and mu-opioid peptides rapidly hyperpolarize GnRH neurons: a cellular mechanism of negative feedback?. Endocrinology. 1995 May 1;136(5):2341–4.

27. Chang RJ, Cook-Andersen H. Disordered follicle development. Molecular and cellular endocrinology. 2013 Jul 5;373(1-2):51–60.

28. Zimmerman Y, Eijkemans MJ, Coelingh Bennink HJ, Blankenstein MA, Fauser BC. The effect of combined oral contraception on testosterone levels in healthy women: a systematic review and meta-analysis. Human reproduction update. 2013 Sep 29;20(1):76–105.

29. Nader S, Diamanti-Kandarakis E. Polycystic ovary syndrome, oral contraceptives and metabolic issues: new perspectives and a unifying hypothesis. Human Reproduction. 2006 Nov 10;22(2):317–22.

30. de Melo AS, dos Reis RM, Ferriani RA, Vieira CS. Hormonal contraception in women with polycystic ovary syndrome: choices, challenges, and noncontraceptive benefits. Open access journal of contraception. 2017;8:13.

31. de Melo AS, dos Reis RM, Ferriani RA, Vieira CS. Hormonal contraception in women with polycystic ovary syndrome: choices, challenges, and noncontraceptive benefits. Open access journal of contraception. 2017;8:13.

32. Welty FK. How do elevated triglycerides and low HDL-cholesterol affect inflammation and atherothrombosis?. Current cardiology reports. 2013 Sep 1;15(9):400.

33. Adams J, Polson DW, Franks S. Prevalence of polycystic ovaries in women with anovulation and idiopathic hirsutism. Br Med J (Clin Res Ed). 1986 Aug 9;293(6543):355–9.

