## supplementary table 1 for "Phenotyping of Indian Polycystic Ovarian Syndrome on the basis of Biochemical Parameters: A retrospective analysis via Hierarchical Cluster Classification"

Supplementary Table 1: Biochemical parameters based on the distribution of LH and FSH ratio

| **Parameters** | **Lab Reference** | **Hormonal Phenotype Group**  **[LH:FSH >2]** | **Metabolic Syndrome Group**  **[LH:FSH =1]** | **P - value** | **P* (adjusted for sample size)** |
| --- | --- | --- | --- | --- | --- |
| No. of patients |  | 21.8 % (54/247) | 78.1% (193/247) |  |  |
| Hormonal Parameters: | | | | | |
| Testosterone (ng/mL) | 0.3 – 0.7 | 0.7 ± 0.33 | 1.08 ± 1.85 | 0.525 | <0.005 |
| Free T3 (pmol/L) | 3 – 8 | 4.28 ± 0.67 | 4.12 ± 0.5 | 0.11 | <0.005 |
| T4 (pmol/L) | 10.6 – 19.4 | 15.66 ± 2.55 | 15.22 ± 2.34 | 0.34 | <0.005 |
| TSH (µIU/mL) | 0.25 – 5.0 | 4.99 ± 11.22 | 3.205 ± 2.67 | 0.12 | <0.001 |
| LH (mIU/mL) | 1.5 – 8 | 11.39 ± 7.37 | 4.2166 ± 2.410 | <0.0001 | <0.0001 |
| FSH (mIU/mL) | 3.9 – 12 | 3.80 ± 2.07 | 4.560 ± 2.157 | <0.05 | <0.001 |
| Estradiol (pg/mL) | <50 | 167.4 ± 200.4 | 64.67 ± 43.06 | <0.0001 | <0.0001 |
| Metabolic Parameters: | | | | | |
| Total cholesterol (mg/dL) | <200 | 147.86 ± 36.11 | 155.43 ± 27.95 | <0.05 | <0.001 |
| Triglycerides (mg/dL) | <150 | 94.11 ± 34.37 | 99.31 ± 37.32 | 0.455 | <0.005 |
| HDL (mg/dL) | >50 | 46 ± 10 | 44.13 ± 9.94 | 0.32 | <0.005 |
| LDL (mg/dL) | <130 | 86.5 ± 26.1 | 91.3 ± 26.8 | 0.35 | <0.05 |
| VLDL (mg/dL) | <40 | 18 ±6.83 | 19.8 ± 7.4 | 0.45 | <0.05 |
| Blood Sugar (mg/dL) | 70 – 110 | 84.22 ± 8.46 | 87.31 ± 13.01 | 0.17 | <0.05 |
| HBA1C (units) | <6% | 5.55 ±0.49 | 5.606 ± 0.357 | 0.53 | <0.05 |
| Insulin Fasting (mIU/mL) | 2 - 25 | 14.23 ± 6.23 | 16.37 ± 6.86 | 0.25 | <0.05 |

Supplementary table 2: Blood glucose index among varies age groups of PCOS patients

| **Age group** | **N** | **Blood sugar**  (mg/dL) | **HBA1C**  (units) | **Insulin Fasting**  (mIU/mL) |
| --- | --- | --- | --- | --- |
| 13 – 18 | 49 | 80.69 ± 7.52  CI ( 77.8 – 83.73)  [69 – 95] | 5.44 ± 0.28  CI (5.2 – 5.5)  [4.8 – 5.8] | 11.37 ± 5.67  CI ( 5.41 – 17.320  [6.3 – 20.3] |
| 19 – 25 | 115 | 85.86 ± 13.26  CI (82.58 – 89.15)  [70 - 157] | 5.617 ± 0.45  CI ( 5.5 – 5.7)  [4.7 – 7.4] | 15.74 ± 7.17  CI ( 13.11 – 18.37)  [5.42 – 32.6] |
| 26 – 30 | 83 | 91 ± 11.5  CI (87.62 – 94.38)  [74 – 132] | 5.6 ± 0.31  CI ( 5.5 – 5.7)  [4.8 – 6.4] | 17.85 ± 5.59  CI ( 15.23 – 20.47)  [8.74 – 29.74] |
| All age group | 247 | 86.53 ± 12.08  CI (84.56 – 88.50)  [69 – 157] | 5.595 ± 0.395  CI ( 5.5 – 5.6 )  [4.7 – 7.4] | 15.69 ± 6.69  CI ( 13.96 – 17.42)  [5.42 – 32.6] |

CI : 95% confidence interval [] –median values

Supplementary Table3 : PCOS Biochemical Parameter across various age groups

| **Age group (years)** | **N** | **Cholestrol**  (mg/dL) | **Triglycerides**  (mg/dL) | **HDL**  (mg/dL) | **LDL**  (mg/dL) | **VDL**  (mg/dL) | **T3**  (pmol/L) | **T4**  (pmol/L) | **TSH**  (µIU/mL) | **LH**  (mIU/mL) | **FSH**  (mIU/mL) | **Estradiol**  (pg/mL) |
| --- | --- | --- | --- | --- | --- | --- | --- | --- | --- | --- | --- | --- |
| Total | 247 | **153.52 ± 30.26**  CI (148.5-158.4)  [ 15 – 244] | **98 ±36.55**  CI (92.04–103.96)  [52 – 252] | **44.6 ± 10**  CI (42.9 -46.2)  [23- 69] | **90.14±26.66**  CI(85.7–94.4)  [33.8 – 178] | **19.62 ± 7.2**  CI(18.4-20.8)  [10 – 50.4] | **4.16 ± 0.55**  CI(4 – 4.25)  [2.6 – 5.9] | **15.33 ± 2.3**  CI(14.9-15.7)  [5.3 – 20.9] | **3.5 ± 5.3**  CI(2.7-4.4)  [0.17 – 60] | **5.9 ± 5.2**  CI(2.7 – 4.4)  [0.1- 31.5) | **4.3 ± 2.15**  CI(4 – 4.7)  [0.1 – 17.5] | **90.5 ± 115.3**  CI(71.7 – 109.3)  [14.34 – 1115.8] |
| 13 - 18 | 49 | **152.96 ± 31.85**  CI (140.10–165.83)  [108 – 218] | **91.54 ± 29.83**  CI (79.49–103.59)  [60 – 203] | **45.23 ± 11.04**  CI (40.7–49.6)  [ 31 – 67] | **89.4 ± 27.4**  CI(78.3–100.4)  [41.6 – 140.2] | **18.38± 6**  CI(15.9–20.5)  [12 – 41] | **4.46 ± 0.65**  CI(4.19-4.73)  [3.63-5.9] | **15.56 ± 1.9**  CI(14.7-16.3)  [12.9 – 19.6] | **3 ± 1.8**  CI(2.3-3.7)  [0.8 – 7.6] | **6.3 ± 6.3**  CI(3.7 – 8.89)  [0.9 – 31.5] | **4.48**  CI(3.8 – 5.16)  [1.4 – 8.18] | **64.5 ± 50.77**  CI(44- 85)  [19.18 – 225.5] |
| 19 – 25 | 115 | **152 ± 26.16**  CI(145.53–158.5)  [ 99 -221] | **99.4 ± 39.34**  CI (89.6 – 109.15)  [52 – 252] | **45.29 ± 9.29**  CI(42.9-47.5)  [23 – 69] | **86.67 ±26.9**  CI(79.9-93.3)  [33.8 – 149.2] | **19.92 ± 7.8**  CI(17.9-21.8)  [11-50.4] | **4.1 ±0.48**  CI(3.98-4.2)  [2.6 – 5.11] | **15.2 ± 2.6**  CI(15.5-15.8)  [5.3-20.9] | **3.7 ± 7.3**  CI(1.9– 5.5)  [0.5 – 60] | **6.5 ± 5.5**  CI(5.3 – 7.8)  [0.9 – 27.15] | **4.11 ± 1.8**  CI( 3.6 – 4.5)  [0.5 – 11.2] | **95.98 ± 98.35**  CI (71.61 – 120.3)  [20.7 – 660.9] |
| 26 – 30 | 83 | **156.15 ± 36.34**  CI ( 145.48–166.82)  15 - 244 | **101.6 ± 36.2**  CI (90.97–112.23)  [52 – 230] | **43.64 ± 10.7**  CI (40.4–46.8)  [23 – 69] | **95 ± 27.2**  CI(87-103)  [44.6 – 178] | **20.34 ± 7.2**  CI(18.2-22.4)  [10 – 46] | **4.09 ±0.56**  CI(3.9-4.2)  [2.9 – 5.8] | **15.4 ± 2.3**  CI(14.5-15.8)  [5.3 – 20.9] | **3.8 ± 3.6**  CI(2.8-4.9)  [0.6 – 17.6) | **5 ± 3.9**  CI(3.9 – 6.2)  [0.1 – 17.6] | **4.2 ± 2**  CI( 3.6 – 4.7)  [0.1 – 9.12] | **100.72 ± 160.5**  CI(53.5-147.8)  [14.3 – 1115] |

CI : 95% confidence interval [] –median values
